# Closing the gaps: Improving physical health diagnosis in the emergency department for patients with mental health conditions

**DOI:** 10.64898/2026.06.05.26354970

**Authors:** Archana Jayaprakash, Elisa Liberati, Rosie Lindsay, Janet Willars, John Gibson, Zoe Fritz, Annabel Price, Thea Hatfield, Natalie Richards, Graham Martin

**Affiliations:** THIS Institute (The Healthcare Improvement Studies Institute), Department of Public Health and Primary Care, University of Cambridge, Cambridge, United Kingdom.; Department of Health Sciences, University of Leicester, Leicester, United Kingdom.; The McPin Foundation, London E2 9DA, United Kingdom.; Cambridgeshire and Peterborough NHS Foundation Trust, Cambridge, United Kingdom.

**Author notes:** Present address: Department of Clinical, Educational & Health Psychology, 1-19 Torrington Place, University College London, London, United Kingdom (Elisa Liberati). Present address: Quantum Consumer Solutions, London, United Kingdom (Thea Hatfield). Present address: Department of Psychology and Human Development, University of East London, London, United Kingdom (Natalie Richards). Corresponding author: Archana Jayaprakash, THIS Institute (The Healthcare Improvement Studies Institute), Department of Public Health and Primary Care, University of Cambridge, Cambridge, United Kingdom.

**Keywords:** mental health, diagnostic inequality, emergency departments

## Abstract

**Objectives:** People with mental health conditions experience increased rates of diagnostic errors and delays in acute treatment. While causes such as diagnostic overshadowing (misattribution of physical symptoms to mental health conditions) are well documented, less attention has been paid to the organisational and structural conditions that shape diagnostic work. This study examines how physical illness is diagnosed in patients with mental health conditions in emergency departments (EDs), with a focus on the structural conditions that enable or constrain safe diagnostic practice.

**Method:** We conducted a multi-site ethnography across three purposively selected EDs in England between April 2023 and April 2024, varying in size, population demographics, and local service configuration. Data were collected through 284 hours of non-participant observation and 20 semi-structured interviews with ED staff.

**Results:** Our analysis identified four recurring structural gaps that shaped the conditions under which physical health diagnosis took place for patients with mental health conditions: a design gap, whereby targets and physical layouts constrained diagnostic reasoning; a preparedness gap, reflecting the lack of structural support to allow staff to act on their existing knowledge and skills; a coordination gap, reflecting fragmented ownership and the challenges of joint assessment across mental and physical healthcare teams; and an expectation gap, whereby unmet need elsewhere in the system increased demand for ED services that were beyond its formal scope. These gaps made diagnostic errors and delay more likely for patients with mental health conditions seeking physical healthcare in the ED.

**Conclusions:** As new dedicated mental health EDs are introduced in England, there is an opportunity to avoid reproducing these structural gaps in new settings. Our study suggests that improving physical healthcare for patients with mental health conditions requires changes to how EDs are designed, resourced and supported, and how they connect with the wider health and care system.

## 1. Introduction

People with mental health conditions face worse physical health outcomes than the general population, including higher rates of chronic disease, increased preventable morbidity, and substantially reduced life expectancy.^1–3^ While these disparities are shaped by a combination of social, behavioural, and treatment-related factors, growing evidence highlights the role of suboptimal physical healthcare, including missed, delayed, or incorrect diagnoses, as a significant contributor.^4–6^

Emergency departments (EDs) are a major point of access into the healthcare system for people living with mental health conditions. They often attend EDs at disproportionately higher rates. For instance, one large survey in England found that, over 12 months, people with only a mental health condition attended the ED twice as often as those without any health condition, whereas those with both a mental and a physical condition attended over four times as often.^7^ As noted by the United Kingdom Royal College of Emergency Medicine, these figures are symptomatic of “a system-wide deficiency or unmet need in physical and mental healthcare as well as social care”.^8^

Despite this significant role that ED plays in the care for patients with mental health conditions, they are not always well equipped to support these patients, including when they present with physical health problems. The ED environment itself can be counter-therapeutic spaces for patients with mental health conditions.^7, 9^ Prior research has identified patterns of stigmatisation in the ED, such as patients being deprioritised or excluded from care pathways (devaluation), or responsibility for their care being deferred or passed on (avoidance).^5, 10^ Recent studies also suggest that patients with mental health conditions often struggle to get their symptoms recognised as legitimate physical health problems, which are more likely to be misattributed or overlooked in the ED, especially when presentations are ambiguous, complex, or atypical.^10, 11^

Recent policy developments in England, such as the introduction of dedicated mental health emergency departments, reflect an increasing recognition that further efforts are needed to provide more integrated physical and mental health care.^12^ Existing studies, however, have largely examined isolated challenges like communication or stigma without fully accounting for the broader organisational and structural context.^6^ Our study addresses this gap. We draw on a multi-site ethnography across three EDs in England to provide insight into the challenges involved in diagnosing physical illnesses in patients with mental health conditions, bringing together interpersonal, organisational, and structural conditions that shape this work. We identify four recurrent, interrelated gaps that shape the ED’s diagnostic work for these patients and argue that improvement requires attention to both clinical practice and the conditions under which it takes place.

## 2. Methods

We draw on a focused sub-sample from a larger multi-site ethnographic study exploring physical healthcare for patients with mental health conditions in EDs. The wider study included data collected from patients, those accompanying them, and staff. In this paper, we focus on staff to examine how the work of diagnosing physical illness is organised and enacted within the ED for patients with mental health conditions. Patient and carer perspectives are reported elsewhere.^10^ For this analysis, we drew on staff interview transcripts and observational data that highlighted staff reasoning, triage decisions, prioritisation logic, coordination between different professionals and teams, diagnostic uncertainty, use of protocols, and navigation of resource constraints within the ED.

We collected data between April 2023 and April 2024 in three EDs in England. The sites were selected purposively based on variations in hospital size, population demographics, and local service configuration. We studied two large urban hospitals (one serving a highly ethnically diverse population) and one medium-sized district hospital.

The study was designed by a multidisciplinary team, including social scientists, a consultant psychiatrist, an acute medicine consultant, and a peer researcher (a researcher with lived experience of mental health care). We convened a group of six expert-by-experience advisors, comprising three service users and three carers. The research team met with the group every six weeks throughout the course of the study, discussing study design, data collection, and emerging analysis.

We obtained Health Research Authority and research ethics committee approval (East of England – Essex Research Ethics Committee on December 22, 2022. IRAS project ID: 265,331) for the study and site-specific approvals for each ED. Written consent was obtained for all interviews, and oral permission for observations.

### 2.2 Data collection

Data were collected through ethnographic observation and semi-structured interviews by five researchers (all social scientists). During observation, we focused on triage, clinical reasoning, team discussions, inter-professional coordination, and how decisions are made under pressure. We captured both verbal and non-verbal interactions in fieldnotes, as well as the contextual and environmental factors shaping care. Following observation, we invited staff to take part in a telephone or on-site interview. Staff were eligible to participate if they had direct experience of assessing or managing physical health concerns in patients with mental health conditions in the ED. They were recruited either when encountered during observational fieldwork, or by researchers liaising directly with staff at each site who helped identify and introduce other eligible members of staff. Qualified doctors, registered nurses, advanced clinical practitioners, ambulance clinicians and paramedics were all eligible for inclusion. Interviews lasted about 40 minutes on average and explored the above topics in depth.

### 2.3 Data analysis

Our analytic approach was iterative, inductive, and grounded in the principles of thematic analysis,^13^ with additional influence from the constant comparative approach.^14^ We began with line-by-line coding of observational fieldnotes and interview transcripts using NVivo 14, identifying recurring patterns and differences across sites and roles. Codes were developed collaboratively, and we organised regular team meetings to discuss and review codes iteratively as the analysis progressed. Through successive rounds of comparison, we grouped initial codes into broader categories that captured recurring structural and organisational issues that influence the diagnosis of physical illness in people with mental health conditions (e.g., “limited training,” “lack of coordination,” “upstream pressures,” “waiting for psych liaison,” “frequent attendance work,” and “time pressure”).

Our analysis revealed that many of these categories pointed to a recurring misalignment between the work required for diagnosis – assessing symptoms, considering competing explanations, coordinating across teams, and deciding next step – and what the ED’s environment enabled or supported clinicians to do. We described these recurring misalignments as ‘gaps.’ We organise our findings around four such gaps, which structure the conditions under which physical health diagnosis takes place in the ED for patients with mental health conditions.

## 3. Results

We conducted 284 hours of observation and 20 staff interviews across three EDs (Table 1). Our analysis identified four recurring forms of misalignment (gaps) that have significant impact on the work of diagnosing physical illness in patients with mental health conditions: the design gap, the preparedness gap, the coordination gap, and the expectation gap. Though analytically distinct, these themes were interconnected in practice and often compounded each other’s effects on staff and patient care. We examine each theme in turn, then provide a synthesis of the improvement suggestions provided by staff.

**Table 1:** Characteristics of sites and interview participants.

|  | Site 1 | Site 2 | Site 3 | Total |
| --- | --- | --- | --- | --- |
| **Site characteristics** | Large hospital | Large hospital in ethnically diverse area | Medium hospital | 3 |
| **Hours of observation** | 90 | 122 | 72 | 284 |
| **Number of interviewees** | 7 | 5 | 8 | 20 |
| **Interviewee roles** | Registrar (4), Foundation year doctor (1), Trainee ACP (1), Clinical fellow (1) | Registrar (2), Nurse (2), Consultant (1) | Foundation year doctor (1), Consultant & lead for frequent attenders (1), ACPs (2), GP Trainee (2), Triage nurse (1), Trust-grade emergency physician (1) |  |

### 3.1 The design gap: how targets, layouts, and workflows shape diagnostic attention in complex ED presentations

Our data revealed a misalignment between the ED’s design and workflow – built for rapid decision-making, throughput, and acute intervention – and the slower, exploratory reasoning that co-occurring mental and physical health problems often demand. Staff repeatedly described how the department’s temporal regime, shaped by rigid performance targets like the 15-minute triage and four-hour discharge rule, left minimal room to explore diagnostic uncertainty, especially in presentations that were complex, ambiguous, or recurrent.

> *“We get told a triage shouldn’t take more than a minute and a half… If you’re rushing through and you just write down, ‘patient has abdo pain,’ the doctors don’t pick up on anything else” (Nurse, Site 2).*

As one registrar (Site 2) noted, the ED ran “at the red line”, with no residual capacity. There was little time or space to pause for reflection or extend an assessment. This left the ED without a buffer against the additional demands that complex or co-occurring presentations involving mental and physical health often created. When cases were ambiguous or required more time than the ED could offer, staff acknowledged that pre-existing psychiatric diagnoses sometimes offered the quickest available explanation. This, they felt, was less a matter of bias than an adaptive response to constraints of time, space, and workload.

> *“This is done in pressured situations, doctors can attribute these kinds of things if the patient has come into the emergency department with repeated attendances of chest pain and he has a history of anxiety and depression, usually he will be labelled as an atypical chest pain patient coming over again and again. But if one or two times, investigations were unremarkable, so if they were to come again with a serious problem, it can be easily missed.” (Clinical fellow, Site 1)*
>
> *“I think if someone presents with a psychiatric or mental health illness that we think would explain x, say, and we’re unable to ascertain another way, it might be easy to attribute it to that instead of trying to find the exact cause.” (Emergency medicine registrar 1, Site 1)*

The observation excerpt in Box 1, while not related to the formation of a diagnosis, further illustrates how these pressures played out in practice, with stretched resources and the absence of residual capacity leading to ad hoc responses, staff frustration, and avoidable risks.

Staff also described how features of the ED’s physical space, such as crowded cubicles, constant foot traffic, high noise levels, and limited privacy, made it difficult to carry out detailed assessments or private conversations about symptoms that might cross mental and physical boundaries. They shaped reasoning by limiting what could be asked, heard, or pursued diagnostically. For instance, clinicians described how they sometimes cut assessments short in shared spaces to avoid making patient uncomfortable or unsafe. Conversely, when quieter and better-resourced spaces were available, clinicians reported greater confidence in holding uncertainty (rather than reaching diagnostic closure prematurely) and attending to both mental and physical aspects of care.

> *“We tried to examine a confused dementia patient, and he picked up the bed remote and flung it at us. In a dedicated mental health [environment], the design put into consideration [of] safety of the medical staff [is greater]. So that gives you more confidence in assessing mental health patients in such places. If you’re concerned about your safety, it makes it more difficult to assess, and so, if they have a problem, you may not be able to identify it.” (Emergency medicine consultant, Site 3)*

These features of ED’s design also led to ‘decision fatigue’.^15^ Across long shifts, the need to provide patients with a high-level diagnostic direction and the sheer volume of such decisions drained clinicians’ capacity for attention and narrowed their diagnostic focus. This reduced their ability to recognise and acknowledge uncertainty, particularly in patients who were difficult to assess or overly familiar from frequent attendances.

> *“The biggest challenges are lack of resources, space, and time. There is a lot of cognitive overload… because on a typical shift, you are making hundreds of decisions in a nine-hour shift and it takes time, patience and a lot of support from the staff, from the senior body, to make sure that [a member of staff] is not fatigued and making a wrong decision.” (Emergency medicine consultant, Site 3)*
>
> *“Typically, it would be a nine-hour shift or ten-and-a-half overnight, which is what I’m doing tonight. And it’s pretty full-on. You know, in a nine-hour shift you get half-an-hour break, otherwise you’re just on the shop floor in an allocated area, seeing potentially undifferentiated patients.” (Foundation year doctor, Site 1)*

#### Box 1. Observation excerpt from Site 3, Day 3

*A patient had come in with an overdose. He was hallucinating, he was trying to get up and walk, but he kept falling over onto the floor, so there was a real risk he would hurt himself. There were about four ambulance crew standing around him, and then another one joined. The security wasn’t coming, so they were talking very loudly about how there’s no system in place. The nurse covering Ambulance Assessment was frustrated that she’d been drafted in from Ambulatory Care, because apparently, they are short-staffed today, and she was really struggling to control the situation, messaging the nurse in charge. The lead consultant was sat at one of the computers but wasn’t interacting with any of this, and it was a very chaotic three quarters of an hour, where people didn’t really know what to do. Security radioed to say they couldn’t come because they were looking after a patient in the Majors Area, where it needed three of the security team, and the nurse in charge was quite frustrated, asking why three were needed when the patient was asleep. The decision was then made to move the patient who had overdosed from Ambulance Assessment to the Majors Area, to be put in a bay next to the patient who the security people were already looking after, so that the same team could cover both. When security eventually arrived, there was an argument between the ambulance staff and security, and later in the Majors Area they moved the bed away and just left the mattress on the floor to minimise the risk of falls*. This excerpt from our observation in Site 3 shows how stretched resources, including time, physical space, and staff, contributed to a loss of coordination during a high-risk, complex presentation. We observed that in the absence of residual capacity the system struggles to absorb even a single complex case without having knock-on effects across the ED. While we did not observe any diagnostic decisions during this episode, the leadership deficit, delayed responses, and improvised solutions pointed to the kind of situation where clinical attention could easily become diffused, and where important assessments might be put off or missed.

### 3.2 The preparedness gap: recognising, assessing, and responding to complex presentations

Across sites, staff described a mismatch between what they were expected to do and what the system enabled or supported when managing presentations involving mental health, especially those that were complex, ambiguous or recurrent. We called this the preparedness gap because it constituted not a lack of awareness but the absence of structures, supports, or routines that would allow staff to act on what they already knew. For instance, many staff recalled receiving mental health training, most often via mandatory e-learning modules that focused on broad topics such as capacity assessment or risk management. However, staff felt that this training was superficial, generic, and disconnected from the clinical realities they routinely encountered.

> *“It’s almost a tick-box exercise… you go through a module online, and it’s like, ‘Okay, I’ve done mental health,’ but then you’re in the ED, and you’re actually with someone who’s very distressed or aggressive and you have no idea what to do.” (Emergency medicine registrar 2, Site 1)*
>
> *“There is diagnostic ambiguity. It’s not clear whether it is a physical health or a mental health problem… even for the seasoned and experienced clinician, it is not as straightforward.” (Emergency medicine registrar, Site 1)*

In describing these cases, staff highlighted the practical competencies they felt were most essential: careful listening, adapting their questions for patients who struggled to describe what was wrong, and staying alert to signs that something potentially serious could be overlooked. Yet clinicians also noted that knowing what to do was not the same as being able to deliver it in practice.

> *“If someone has delirium we know side-rooms and one-to-one supervision help, but you ask for that and you’re met with a grim laugh.” (Emergency medicine registrar, Site 2)*
>
> *“Sometimes that 15 minutes isn’t enough, especially when people with mental-health problems are low, withdrawn or overdosed. It just takes longer to get a history… I’m able to picture the things that need doing. We’ve got the structures and the training, but some days you’re stuck with a patient because no-one else can supervise, and everything backs up.” (Nurse 1, Site 2)*

One recurrent example in staff’s accounts was the challenge of not letting previous psychiatric labels unduly influence their decision-making. Staff emphasised that the ability to hold competing explanations in mind and weigh new symptoms and old diagnoses without jumping to conclusions was a core aspect of preparedness for complex presentations. However, they found this diagnostic openness difficult to sustain in a setting that prioritised throughput and had limited opportunities for reflection or specialist input from other teams. One emergency medicine registrar (Site 1) described how, as workload intensified, their ability to *“see the wood for the trees”* (hold the broader picture in view) diminished, giving rise to a form of *“tunnel vision”*.

Participants reported relying heavily on informal learning, especially from senior or more experienced colleagues. While this peer-to-peer learning was valued and acknowledged as an important feature of medical training, it was also noted that more standardised and structured teaching around physical health diagnosis in people with mental health conditions was lacking, potentially contributing to variability in care, and leaving staff concerned about gaps in preparedness, especially for presentations involving mental health.

> *“Each person who’s investigating… you get varied qualities [because] people [have] different other specialty experiences and different training backgrounds. And that’s an inconsistency that probably would need to be fixed.” (Emergency medicine registrar, Site 3)*

These accounts also highlighted the absence of formal systems to consolidate and share learning around mental healthcare in the ED. Without structured guidance, clinicians were often left to ‘re-invent the wheel’ with each shift. As one clinician reflected, protocols were sometimes only introduced reactively, after a serious incident:

> *“Previously one of the doctors had [a] serious incident… the patient was very agitated. They tranquilised and unfortunately the patient went into cardiac arrest… So that’s why they were looking at the protocol in the department on how to intervene in mental health patients so that they can be properly assessed. Sometimes we only develop local guidelines after something goes wrong… but that shouldn’t be the case.” (Emergency medicine registrar, Site 2)*

### 3.3 The coordination gap: boundaries, ownership, and handover in the ED

Staff described how the day-to-day work of coordinating care for patients with both mental and physical health needs was disrupted by the way people, teams, routines, and organisational boundaries interacted, or failed to cohere, within the ED. One triage nurse (Site 2) noted that it fell to ED staff to assess not just the severity but the nature of patients’ symptoms (whether physical health– or mental health-related), often based on the “presenting problem” identified at triage. If the problem was straightforwardly physical, ED staff would carry out investigations, treat the patient, or refer them to other specialty teams. However, when mental health concerns were present (either as the presenting problem or alongside physical symptoms), it generated significant ‘articulation work’^16^ for ED clinicians: the often-invisible, ongoing effort required to coordinate tasks and actors.

> *“I really understand that every specialty is really busy, and they don’t want to see anything that is not for them, but at the same time it is the A&E doctor’s job to do this big almost sorting operation.” (Advanced care practitioner, Site 3)*

A key example of articulation work^16^ was “medical clearance”, a process through which any underlying physiological causes were ruled out before a patient was transferred for psychiatric assessment.^17^ Some participants described medical clearance as a necessary step and “good practice” before any mental health issues were considered, with one advanced care practitioner (Site 3) explaining that clinicians had to be *“absolutely sure there is no physical cause before psychiatry will see them.”* However, others called for more collaborative models where both mental and physical health issues were considered together, especially when mental health problems shaped the way physical symptoms appeared, or when mental health conditions, such as anxiety or phobias, made it hard for patients to explain their symptoms or take part in diagnostic tests. Without the option for early, joint assessment, ED clinicians had to make sense of these complex presentations on their own, sometimes struggling to decide whether further tests were necessary or when to involve psychiatry.

> *“You need someone, often it’s a multidisciplinary team, including a psychiatrist in emergency medicine, who will be able to say with much more authority [that] doing blood tests is not going to be of benefit, for example.” (Emergency medicine registrar 1, Site 1)*
>
> *“They need to be ‘medically fit’ before we do a mental health review. Whereas the Royal College of Psychiatry is quite clear that shouldn’t stop them reviewing. I completely disagree with it. Because there’s so much of overlap and overlay.” (Emergency medicine registrar, Site 3)*

Clinicians also recognised that medical clearance could encourage teams to focus narrowly on their own defined tasks, inappropriately segmenting physical and mental health diagnoses in ways that did not reflect their interdependence. For example, once medical clearance was completed and a psychiatric referral made, there was a tendency to relinquish ownership of the case, even when ambiguity remained, increasing the risk of premature diagnostic closure.^18^

> *“There’s a subconscious tendency to do a ‘medical clearance’ then refer on to psych liaison, which has the potential to cause serious harm.” (Emergency medicine registrar, Site 3)*
>
> *“Bias plays a role – surgeons sometimes think, ‘Oh it’s probably nothing’, if the patient has anxiety. But if they [need] investigations like MRI [it] can be hard because many patients are claustrophobic, and that makes it hard to get the scan done.” (General surgery registrar, Site 2)*

These challenges were further reinforced by the organisation of services. Psychiatric liaison teams were often based in separate buildings, short-staffed, or only available during certain hours. Staff noted how these constraints sometimes delayed specialist input and made it harder for clinicians to work together as a single team, exacerbating fragmentation.

> *“The mental health team work in another building; they come over to EDU/ED but have no base here. They see the patient, discuss with us, then we request scans or bloods. It works, but it’s a lot of back-and-forth.” (Advanced care practitioner trainee, Site 1)*
>
> *“Liaison Psychiatry are excellent in hours; out-of-hours it’s difficult* – *you’re phoning another hospital who can’t get here quickly. Specialist input is very time-dependent.” (Emergency medicine registrar 2, Site 1)*

### 3.4 The expectation gap: unmet needs, frequent attendance, and the toll on ED staff

The ED’s open door meant it often saw patients whose needs fell outside its intended remit. When primary care, community mental health, social care, or crisis teams could not respond (due to long waits, strict eligibility thresholds, or limited out-of-hours cover), patients were redirected or self-referred to the ED. As a result, the ED absorbed longstanding unmet needs and complex issues spanning social, mental, and physical domains, many requiring time, coordination, and specialist input. This created an expectation gap: a mismatch between what the ED was designed and resourced to do and the work it was routinely required to absorb.

> *“We’ll get family that’ll come in and say, we’re not getting anywhere with the mental health support team, you need to do something… we’re worried if this person goes home what’s going to happen.” (Nurse 1, Site 2)*
>
> *“I’ve had handovers where the paramedics have told me that there’s no food in the cupboards. Sometimes they come in because they’re living in a cold apartment. It’s not necessarily that they are medically unwell, but because their social situation is not great, we would admit them for a period until their social situation was taken care of by external services.” (Emergency medicine consultant, Site 3)*

Staff described how delays or failures in upstream services brought new forms of work into the ED. We refer to this as emergent work: unplanned tasks that arrived in the ED because upstream services could not respond, and the ED staff had to absorb them immediately. Staff explained that this work placed significant strain on the ED. It required them to manage a range of ongoing needs, including problems that would more appropriately be addressed by other services. This further stretched already limited ED capacity, narrowing the time and attention available for diagnostic reasoning. Over time, the persistent presence of emergent work also eroded morale, as staff were expected to deliver care without the means to do so.

> *“As a clinician in the NHS, your hands are tied because there’s actually nothing much you can offer these patients. Even if you wanted to admit them just for pain relief, it’s difficult to justify in the current climate because we don’t have the resource, we don’t have beds.” (Foundation year doctor, Site 3)*

Staff sometimes coped with the demands of emergent work through avoidance tactics^5^ such as distancing themselves from patients, deferring engagement, or passing responsibility to others. Staff described how these responses were particularly pronounced for presentations involving mental health. For example, some clinicians described instances where patients with a known mental health history were avoided, as they were seen as time– and resource-intensive. This included anticipating the need for close supervision, liaison calls, or longer assessment time.

> *“If [this patient] needs one-to-one supervision, that’s a member of staff you’ve lost there, causing disruption to the department.” (Trust grade emergency physician, Site 3)*
>
> *“These histories take longer, people tend to avoid them, because I guess you’re under pressure to see a certain target number of patients. There’s a lot of resource expended.” (Foundation year doctor, Site 3)*

As a result, their physical health concerns were more likely to be downplayed or inadequately investigated: an experience of devaluation,^5^ whereby certain patients or problems come to be seen as less deserving of attention or resources. Devaluation was shaped both by the pressures of emergent work and by prevailing attitudes about which cases belonged in the ED.

Frequent attenders (patients who used the ED frequently, often with medically unexplained symptoms) offered a particularly clear example of devaluation in practice. Staff said that many of these patients arrived with both physical complaints and a recorded mental-health history, so their assessment was expected to take longer and to involve more staff. Over time, clinicians said, this pattern began to feel cyclical and increasingly futile, leading them to question whether another set of tests or another brief admission would change anything.

> *“These attendances may be of no or little medical therapeutic benefit and they fall into a challenging pattern of behaviour that isn’t doing the patient any good and definitely isn’t doing us any good.”* (Emergency medicine registrar 1, Site 1)

For staff, the repeated presentation of these patients created a dilemma. Investigating too little risked missing serious illness. Investigating too much would expend limited resources and might reinforce patterns of repeated, unresolved care. Many staff expressed uncertainty about how best to respond.

> *“A lot of the time these patients are actually over-investigated. Whether those investigations are appropriate, I don’t know* – *I don’t have evidence for it. But I’ve seen cases where a patient undergoes multiple investigations, gets referred on, has even more tests… and you wonder, was that the right decision? Maybe not. She didn’t need that overnight scan. Someone else might have needed that slot more.” (General surgery registrar, Site 2)*
>
> *“If they’re really anxious, or insistent that they want this particular investigation… I’m not going to get in the way of that because it isn’t worth it to me to spare them that radiation dose if they’re worried about it.” (Emergency medicine registrar 1, Site 1)*

Clinicians thus faced a tension between defensive medicine, departmental capacity, and a growing awareness of the risks involved in over-investigation, both for the individual exposed to radiation unnecessarily and for others who might be more in need of tests.

### 3.5 Suggestions for improvements

Across the sites, staff suggested improvements to address some of the challenges described above (Table 2). These included changes to time and space for assessment, more practical and locally relevant training, clearer referral pathways and access to specialist input, better documentation to support continuity of care, and stronger support for staff wellbeing.

**Table 2:** Summary of key challenges and staff suggested improvements.

|  | **Key challenges identified by staff** | **Staff-identified improvements** | **Illustrative data** |
| --- | --- | --- | --- |
| 1 | Focus on time targets and quick throughput increases the risk of misattribution of physical health symptoms to mental health diagnosis, leading to missed physical illnesses. | Improve documentations of past assessments; enhance staff training on unconscious bias; allocate protected time for training; reduce workload strain so staff have more time on assessments. | *"There are trainings in place, but they come bundled up with a lot of other things which are not as interesting, and equally there's no real time allocation to do them... Training in not using unconscious bias is probably a really good thing."* (Triage Nurse, Site 3) |
| 2 | Policy targets such as 15 minutes triage and discharge within four hours of arrival in an ED do not account for the realities of EDs, often pushing rushed assessments that disadvantage patients with complex and overlapping needs. | Increase flexibility in triage timing for complex cases; implement longer assessment slots for mental health patients. | *"There's a 15-minute window for paramedics to get a patient from the ambulance in and handover by the bedside, and they want us to...it's all about numbers and [throughput] on seats and red tape, so there is...yeah, there's a...you should be able to do a triage in 90 seconds and have that...and then be calling the next patient in."* (Nurse, Site 2) |
| 3 | The ED's physical environment is poorly suited to patients with mental health needs regardless of the | Designate quiet spaces for mental health patients; adapt ED layouts to better accommodate needs of patients with mental health needs. | *"It'd be nice to have a kind of dedicated route where patient presents with mental health, they're streamed to a quieter environment where they'd be looked after from that perspective by a specialist clinician, but we just don't have the ground space* |
|  | reason for their attendance. |  | *and staffing to do that, really.” (Trainee ACP, Site 1)* |
| 4 | Limited mental health training is available for ED staff. | Increase mandatory mental health training for ED staff, introduce psychiatry rotation for ED trainees, and make training more practical by focusing on managing crises (e.g., de-escalation. Restraint techniques), handling complex presentations involving overlapping mental and physical health needs, and engaging with frequent attenders. | *“When we’re being asked to manage this case in the first instance, with very little training, it’s quite hard because they have lots of training in cardiology and respiratory, and everyone does their rotations through these jobs, but there’s plenty of people that have never done a rotation through psychiatry, so I think that makes management of these patients even harder.” (Foundation year doctor, Site 1)* |
| 5 | Lack of clear referral pathways for presentations involving mental health, both within ED and to external services, leads to delays and gap in care. | Standardise clear referral pathways both within ED and to external services; more awareness about different services and referral pathways. | *“Sometimes I struggle with it, what would be the right health service they should actually go to? Psychiatric would not accept these kinds of patients right away in the ED, so the best idea is to when they come to you, just reassure them about their symptoms, say there is nothing going on... I don’t know the exact pathway how and where they need to be referred.” (Emergency medicine registrar, Site 2)* |
| 6 | Lack of primary care appointments, community mental | Expand access to psychiatric beds and community crisis teams; | *“A lot of the history we see is that I haven’t been to my GP for the last year, I’ve tried to get appointments, I’ve tried to access this. The wait* |
|  | health services, and psychiatric beds leads to prolonged ED stays. | improve coordination between ED and primary care. | *times for mental health services is astronomical. The cost for private services is very much not something that many of the people we see can afford to access." (Advanced clinical practitioner, Site 3)* |
| 7 | Patients with both physical and mental health needs often 'fall through the cracks' without joint assessments. | Introduce joint physical-mental health assessment within the ED; increase availability of specialist input. | *"The key would be more specialist opinion help, informally or formally. It's incredibly useful when patients come with a nurse who knows them and can provide exact events leading up to their visit. Having a contact number for their named consultant is also helpful but doesn't always happen. We often worry about capacity, consent, and ensuring we don't hinder important mental health treatments, especially when patients are on multiple antipsychotics, avoiding harmful interactions." (Medical registrar, Site 1)* |
| 8 | Frequent attenders risk being dismissed as "repeat cases", potentially exposing them to missed diagnosis of emerging physical health problems, or over-investigation. | Document each visit, reason for attendance, and outcome; engage multidisciplinary teams or liaison service early on. | *"There should be very clear documentation to help us out from previous attendances, on exactly how they present, what about them could typically be [driving repeated attendance], de-escalation, or things which would exacerbate the situation. And you need someone, often it's been multidisciplinary team, including a psychiatrist in emergency medicine, who will be able to say with much more authority [that] doing blood tests is not going to be of benefit, for example." (Emergency medicine registrar, Site 1)* |
| 9 | Cognitive burden and moral distress for ED | Increase staff supervision and support; implement burnout prevention | *"The knowledge and training is pretty good. I think the resources to execute it is often lacking. And that's a pretty well-established source of burnout,* |
|  | staff increase with their workloads. | strategies; address moral distress with peer support. | *isn't it? That you can...you're witnessing some of that need on a regular basis and you're trying to practise to the standard which you've been trained to and you're forced to compromise your standards of delivery." (Emergency medicine registrar, Site 2)* |

## 4. Discussion and conclusion

Prior research has shown that patients with mental health diagnoses experience increased rates of diagnostic delay and error in acute treatment.^11, 19^ Many studies attribute this disparity to clinicians’ individual biases.^5, 20^ This attribution has been beneficial in part and has led to increased awareness and mental health training. However, it also risks implying that the problem lies solely with individual clinicians, and therefore the solutions lie there too.

Our study complicates this notion. Across the three EDs we studied, we found that clinicians were largely aware of diagnostic overshadowing, concerned about it, and actively trying to avoid it. However, our study shows that awareness alone was insufficient without the organisational conditions that allow clinicians to translate their awareness into reliably safe diagnostic practices for patients with mental health conditions. We identify four interrelated gaps – design, preparedness, coordination, and expectation – that help explain this disconnect. We argue that these gaps are structural features of how the ED is resourced and positioned within the wider healthcare system and produce conditions in which diagnostic problems are more likely for patients with mental health conditions.

The design gap shows how EDs are structured around a logic of rapid resolution. This is very evident in the significant emphasis placed on time targets and throughput metrics such as the four-hour standard, requiring that patients are seen and either admitted, transferred, or discharged within four hours of arrival. But as Bevan and Hood have observed, “hitting the target and missing the point”^21^ can become routine when such metrics dominate practice. Our findings showed how these targets and metrics can particularly disadvantage patients with mental health conditions. Staff emphasised that time pressures left little opportunity to understand symptoms that were complex, fluctuating, or difficult to separate from mental health history, increasing the risk that physical problems are overlooked or attributed to mental health diagnoses.

Many staff also described experiencing decision fatigue. They saw this as a direct consequence of the ED’s design, characterised by overcrowded cubicles, constant foot traffic, high noise levels, and lack of privacy, making it harder for clinicians to gather information, communicate sensitively, and build trust, especially with patients experiencing distress, anxiety, or confusion. Consistent with the dual-process typology^22, 23^, staff reported defaulting under these pressures to Type 1 reasoning – that is, rapid pattern recognition that can sometime lead to premature diagnostic closure. A recorded mental health diagnosis could thus become the organising explanation for an attendance. Though efficient for straightforward presentations, clinicians recognised that this form of reasoning becomes risky when mental health histories complicated assessment, requiring slower, analytic (Type 2) reasoning.^22, 23^

We argue that underlying these pressures is a lack of slack^24^: available spare resources, of any sort (time, space, staff, etc.), which can be called on in times of need” to absorb complexity and uncertainty. The tendency to default to familiar explanations and to reach premature closure may be understood in part as a pragmatic adaptation to an environment with insufficient slack or residual capacity. Staff accounts consistently emphasised that managing these presentations safely requires time to extend assessment when required, opportunity to revisit earlier decisions, access to senior colleague to discuss uncertainties, and capacity to maintain diagnostic openness without reaching for premature conclusions.

Our findings do not contradict existing evidence showing how individual clinicians’ bias and knowledge of mental health influence diagnostic decision making in high-pressured environments like EDs.^5, 20^ Rather, our notion of the preparedness gap complements these analyses by helping to explain the conditions that perpetuate these deficits. Most staff said they had received some form of mental health training, but many still felt underprepared when dealing with patients whose needs spanned both mental and physical health. Staff emphasised that, in the ED, preparedness for complex presentations involving mental health depends on immediate senior support, fast access to information, access to liaison psychiatry, adequate staffing, and the time to discuss cases with a senior or more experienced colleague. When that is missing, staff fall back on ad hoc workarounds and cannot put their training to full use, illustrating the difference between individual competence and the collective competence that emerges only when a team can pool and coordinate its expertise.^25^

This preference for joint assessment and coordinated response was clear across the three sites. However, the way care is organised in the ED makes this difficult to achieve in practice: who is responsible for what, and when, is often defined in ways that are not designed for presentations where mental health histories complicate physical health assessments. It is in navigating these boundaries that the coordination gap becomes most visible. Medical clearance offers an illustrative example. While some participants regarded medical clearance as good practice, others acknowledged that it could harden the boundary between ED clinicians and psychiatric liaison teams when shared reasoning is most needed. When early joint assessment is absent, ED clinicians alone must determine whether symptoms reflect physical illness, mental health issues, or both: a form of articulation work^16^ that is time-consuming, cognitively demanding, and not always delivering the best experience for patients.

The ED’s functioning is also shaped by how it connects with other upstream and downstream services. Its ‘open door’ means that it often serves as a default service for people who are unable to access primary care, mental health services, or community support, a pattern sometimes referred to as ‘failure demand’.^26^ While failure demand helps understand why patients arrive in the ED, it does not explain what happens once they do. Our analysis shifts the focus from demand itself to the expectation gap it produces, and the work it generates. In the ED, this shows up as a steady stream of emergent work: unplanned tasks that are outside the ED’s remit but still have to be managed on arrival, even if sub-optimally. This work typically does not fit easily within the training, staffing, or routines of the ED, such that staff have to adapt, coordinate, and find solutions on the spot, acting as shock absorbers^27^ for gaps elsewhere in the system.

To manage the disruptions that emergent work creates within the ED, staff developed local adaptations that helped them absorb the displacement of work from elsewhere. However, prior studies show that even if they work in the short term, such local adaptations come at a cost: workarounds become routine and mask underlying problems, while clinicians’ attention is repeatedly drawn to the most interruptive demands, reducing the decision time available for complex presentations.^28, 29^ We found the same pattern. Staff acknowledged the impact of emergent work on patients already at greater risk of facing diagnostic inequalities in the ED. They described how patients with known mental health histories or frequent attendances were often seen primarily in terms of the resources they required, such as time, staffing, emotional effort, and coordination. As a result, those already perceived as resource-intensive were at greater risk of devaluation or avoidance, forms of structural stigma well documented elsewhere.^5, 20^

This has important implications for staff wellbeing. Clinicians described experiencing moral distress when they could not provide the standard of care they regarded as necessary. As prior research shows, moral distress among emergency clinicians increases the risk of staff leaving the workforce and of diagnostic errors.^30^ We thereby show that efforts to improve diagnosis cannot ignore the conditions of the work in the ED, including the emergent work generated by failures elsewhere and its downstream effects on staff capacity, wellbeing, and the time and attention available for complex presentations involving mental health.

Across the three sites, staff made suggestions for improving physical health diagnosis (and more broadly physical healthcare) for people with mental health conditions in the ED (see Table 2), setting out ideas to address the design gap in particular. Specifically, suggestions such as greater flexibility during triage for complex presentations involving mental health, quieter and safe spaces for assessment, and more support from liaison teams, were highlighted as possible ways to support more detailed assessments. Even when some of these recommendations from staff represent more modest adaptations possible within the constraints of existing resources, others point to broader capacity issues (such as workforce shortage) that may be difficult to address through local service redesign alone. Further research is required to determine the most effective ways of improving diagnostic safety for patients with mental health conditions seeking physical healthcare in the ED, and to understand what improvements may be possible to achieve through local organisational changes.

Our study has some limitations. First, as an ethnographic study based on staff accounts and observations, we did not include a comparator such as people without mental health conditions or sites with different liaison psychiatry models, nor did we measure diagnostic outcomes. Some of the challenges we describe, such as time pressure or limited space, extended to all patients, even though their consequences seemed to be magnified for those with a mental health condition. Second, although the larger study included patient and carer perspectives, this paper is based on the work of staff, as evidenced in interviews and ethnographic observations. Finally, even though we observed three EDs across shifts, our fieldwork may not capture rare high-acuity or out-of-hours events, and an observer effect (whereby the presence of a researcher influenced the behaviour of staff) cannot be ruled out. We have tried to overcome these challenges through prolonged engagement, triangulation between data types (observation and interviews) and across staff roles, and discussions with our multidisciplinary team, which included a consultant psychiatrist, an acute medicine consultant, and a peer researcher with lived experience of mental health care.

In England, as new dedicated mental health EDs are introduced,^12^ there is an opportunity not only to redesign services, but also to reconsider the organisational assumptions that shape diagnostic work. The four gaps identified in our study foreground the need for structural changes that explicitly accommodate uncertainty, make space for slower reasoning, and support the relational work of holding cases open across professional and organisational boundaries. Crucially, because these are gaps produced by the affordances and constraints built into how space, time, roles, and responsibilities are configured, they are also modifiable, as evidenced by the suggestions staff themselves proposed. Acting on these suggestions provides a potential way to address the diagnostic inequalities experienced by patients with mental health conditions in acute care settings, as well as the challenges that staff face in delivering care in current EDs and in the new mental health EDs now taking shape.

## Acknowledgements

We are grateful to our expert-by-experience consultation group whose feedback shaped the study from its conception. We also thank Annabelle Olsson for her support with data collection and the staff at our study sites for facilitating fieldwork.

## Ethical approval and informed consent statements

This study had received ethics approval by the East of England – Essex Research Ethics Committee on December 22, 2022. IRAS project ID: 265,331. All participants provided informed consent.

## Data availability statement

Due to the nature of the study and the risk of deanonymisation, data are not available for this study.

